# SARS-CoV-2 virus and antibodies in front-line Health Care Workers in an acute hospital in London: preliminary results from a longitudinal study

**DOI:** 10.1101/2020.06.08.20120584

**Authors:** Catherine F Houlihan, Nina Vora, Thomas Byrne, Dan Lewer, Judith Heaney, David A. Moore, Rebecca Matthews, Sajida Adam, Louise Enfield, Abigail Severn, Angela McBride, Moira Spyer, Rupert Beale, Peter Cherepanov, Kathleen Gaertner, Sarah Edwards, Maryam Shahmanesh, Kevin Ng, Nikhil Faulkner, Georgina Cornish, Naomi Walker, Susan Michie, Ed Manley, Fabiana Lorencatto, Richard Gilson, Sonia Gandhi, Steve Gamblin, George Kassiotis, Laura E McCoy, Charles Swanton, Andrew Hayward, Eleni Nastouli

## Abstract

**Background:** Although SARS-CoV-2 infection in Healthcare Workers (HCWs) is a public health concern, there is little description of their longitudinal antibody response in the presence or absence of SARS-CoV-2 and symptoms. We followed HCWs in an acute London hospital to measure seroconversion and RNA detection at the peak of the pandemic.

**Methods:** We enrolled 200 patient-facing HCWs between 26 March and 8 April 2020 and collected twice-weekly self-administered nose and throat swabs, symptom data and monthly blood samples. Swabs were tested for SARS-CoV-2 by PCR, and serum for antibodies to spike protein by ELISA and flow cytometry.

**Findings:** During the first month, 42/200 (21%) HCWs were PCR positive in at least one nose and throat swab. Only 8/42 HCW (19%) who were PCR positive during the study period had symptoms that met current case definition. Of 181 HCWs who provided enrollment and follow-up blood samples, 82/181 (45.3%) were seropositive. In 33 HCWs who had positive serology at baseline but were PCR negative, 32 remained PCR negative. One HCW had a PCR positive swab six days after enrollment, likely representing waning infection.

**Conclusion:** The high seropositivity and RNA detection in these front-line HCWs brings policies to protect staff and patients into acute focus. Our findings have implications for planning for the ‘second wave’ and for vaccination campaigns in similar settings. The evidence of asymptomatic SARS-CoV-2 infection indicates that asymptomatic HCW surveillance is essential, while our study sets the foundations to answer pertinent questions around the duration of protective immune response and the risk of re-infection.

## Introduction

Health Care Workers (HCWs) were shown to be at risk of infection during the 2003 outbreak of Severe Acute Respiratory Syndrome Coronavirus (SARS-CoV); 21% of worldwide cases were thought to have occurred in HCWs, and infection acquisition in hospitals was clearly demonstrated (1,2). In Wuhan, where SARS-CoV-2 emerged the incidence of SARS-CoV-2 was higher in HCWs than the general public (3). While some studies have suggested a higher seroprevalence in HCWs, there remains a critical lack of data on how SARS-CoV-2 serology changes over time, or on the protective effect of seropositivity against reinfection in high exposure settings (4)

Occurrence of asymptomatic infection in HCWs(5–7) and onward transmission of SARS-CoV-2 has been demonstrated (8,9). However, whether asymptomatic infection leads to detectable, and longitudinally stable antibody titres and function is unknown.

To curb the spread of the pandemic, several countries including the UK have introduced restrictions in social mixing. Despite these measures, health care facilities have been implicated as areas of ongoing virus transmission and UK recommendations were thus altered to mandate personal protective equipment (PPE) for all patient contact (10). After the lockdown, a reduction in new COVID-19 cases was seen. However the possible effect of alterations in PPE recommendations on the incidence of infection in HCWs has only been reported in one small study (7).

Despite the clear risk to HCWs and potential risk to patients, there has been a lack of longitudinal data on seroconversion and evidence of SARS-CoV-2 RNA in seropositive and seronegative individuals. To address this, we carried out “SARS-CoV-2 Acquisition in Frontline Healthcare Workers - Evaluation to inform Response (SAFER)”, a prospective cohort study in high risk front line HCWs with regular serology and self-administered swabs, in acute NHS hospital trusts in London and Liverpool. We aim to generate essential data on risk of infection in front-line HCWs, the potential for asymptomatic transmission and the possibility of re-infection. Here we present data from the first complete month of the study in London.

## Methods

### Enrolment and follow-up

We enrolled HCWs at University College London Hospitals between 26th March and 8th April 2020. Eligibility criteria were being asymptomatic at time of enrolment and working in one of five clinical areas: Accident and Emergency (A&E), acute medical admissions (AMU), COVID-19 cohort wards, Intensive Care Unit (ICU) and haematology wards. We invited HCWs to discuss the study with a research team member before or after hand-over meetings, or in the wards or rest areas without interruption to patient care. No incentives were offered for enrollment. With consent, we conducted baseline interviews including demographic data, self-reported comorbidities, and information on patient contact and aerosol generating procedures (AGPs). Participants then performed a single observed self-administered nylon flocked combined nose and throat swab with instructions. The research team collected 10mls of blood in serum separating tubes (SST), transported daily to the laboratory for processing and storage at −80°C.

Participants submitted follow-up self-collected swabs and symptom questionnaires including patient contact and AGPs twice per week for 3 months. Participants also collected samples at home when not on shift, stored in a refrigerator and submitted on return in <7 days… Text messages and face to face reminders were used. HCWs were informed that results would be provided after one month due to PCR testing capacity limitations.

The study protocol was approved by the NHS Health Research Authority (ref 20/SC/0147) on 26 March 2020. ethical oversight was provided by the South Central Berkshire Research Ethics Committee

### Laboratory testing

As part of the pandemic response, UCLH formed a partnership with the Francis Crick Institute (the Crick-COVID-Consortium) to increase diagnostic capacity and swabs were tested for SARS-CoV-2 RNA by PCR (11). Briefly, 100uL of thawed viral transport medium was inactivated by mixing with 1ml of 5M guanidine thiocyanate L6 virus inactivation buffer. 150uL of this inactivated lysate was used for RNA extraction using a binding buffer and silica-coated magnetic beads (GBiosciences) and guanidine hydrochloride solution. 10 μL of extracted RNA underwent real-time reverse transcriptase PCR on the ABI QuantStudio 3 using primers and probes from the commercial BGI kit which detects the ORF1ab region of SARS-CoV-2. The kit contains a human beta-actin internal control that confirms swab adequacy .All samples were tested in duplicate on separate 96 well plates, with each run containing a blank negative control and positive external control, results reported based on the detection of the internal control and SARS-CoV-2 for each sample, as per the manufacturer’s instructions.

If two results on the duplicate runs were discordant, i.e. the first test result was negative (Cycle threshold - CT 0 or CT >37) and repeat result was weakly positive (CT >35.0, <37.0) then it was reported as negative. If two samples gave discordant results where one was strongly positive (CT<35) and the other negative (CT=0 or CT>37.0), the sample was repeated a third time using a different assay. The second assay was an in-house rt-PCR which detects the nucleocapsid (N) gene of SARS-CoV-2 on the Hologic Panther Fusion that includes Hologic internal control primers and probes (12).

Serum was tested using in-house flow cytometry and ELISA assays developed as part of the Crick-COVID-Consortium (13). Briefly, flow cytometry used HEK293T cells which expressed wild-type SARS-CoV-2 Spike protein bound to a 96-well plate. Cells were incubated with participant serum diluted 1:50 with phosphate buffered saline (PBS) and washed with FACS buffer before being stained with anti-IgG, M and A. Plates were read on a Ze5 (Bio-Rad) running Bio-Rad Everest software v2.4. Analysis was carried out using FlowJo vlO (Tree Star Inc.). A flow assay result was considered positive if the number of IgM+IgA+IgG+ cells was greater than or equal to 28%.

For the ELISAs, 96-well plates were coated with recombinant S1 protein (UniProt ID P59594, residues 1-530) and blocked using a blocking buffer. Participant serum was diluted 1:50 with blocking buffer and 50 μL added to the plate. Plates were incubated for 2 hours, washed 4 times with PBS-0.01% Tween and incubated with alkaline phosphatase-conjugated goat anti-human IgG for 1 hour before a further 6 washes with PBS-0.01% Tween. Plates were read after the addition of alkaline phosphatase substrate for 30 min. Optical densities (OD) were read at 405 nm on a microplate reader (Tecan). Each plate included a positive and negative control. Plate quality control required the average positive control OD exceeded 2 and no individual negative control OD to exceed 0.25. The lower cut-off was set at 0.4 as this was >4-fold above the average of wells containing secondary antibody only across all plates. The upper cut-off was established by prior evaluation with serum from SARS-CoV-2 PCR positive patients and pre-pandemic negative controls. Results were then read as follows; <0.4 was negative, 0.4 to <0.9 was indeterminate and greater than or equal to 0.9 was positive.

The composite serology results were classified as positive if the flow assay was positive irrespective of ELISA, indeterminate if the flow assay was negative but ELISA strongly positive (>0.9) and negative if the flow assay was negative and ELISA <0.4 or between 0.4 and 0.9 OD.

### Data management and analysis

Data from the questionnaires were double-entered onto a REDCap database(14) and analysed using R v3.5.1 and STATA V15. We described the baseline demographics and the cumulative outcome of ever having had a positive swab with SARS-CoV-2 or having a composite positive serology result. Positive results were described within the demographic and exposure variables using fisher’s exact tests.

Sample size calculations were based on the precision around the cumulative proportion of staff expected to acquire SARS-CoV-2 over the study period. The study was designed to enrol 100 HCWs which would allow a precision of +/- 10% to detect a prevalence of 50% with 95% confidence. The increase to 200 was due to a recognition of enrollment demand and the award of funding.

The data for this interim analysis were extracted on 10^th^ May 2020. Symptom data reported at the time of the swabs was summarised. Symptoms recorded, which were compatible with a diagnosis of SARS-CoV-2 according to Public Health England were reported fever, new continuous cough or a new alteration in sense of taste or smell (15). In order to construct an epidemiological curve, we included data on the number of new diagnoses seen per day in London (**https://coronavirus.data.gov.uk**) and individuals were plotted at their first SARS-CoV-2 PCR-positive date.

## Results

We enrolled 200 HCWs between 26th March and 8th April 2020. The median age was 34 years (inter quartile range (IQR) 29-44) (Table 1). We enrolled 82 nurses, 72 doctors, 12 physiotherapists, 11 healthcare assistants, 10 staff working in catering (delivering food) or administration (ward clerks), 3 porters, and 3 staff working in sanitation or housekeeping. Participants were equally distributed across selected areas. The mean number of swabs submitted was 8.5 of 10.3 expected, per participant; 146 participants (73%) submitted at least 1 swab per week.

**Table 1.**
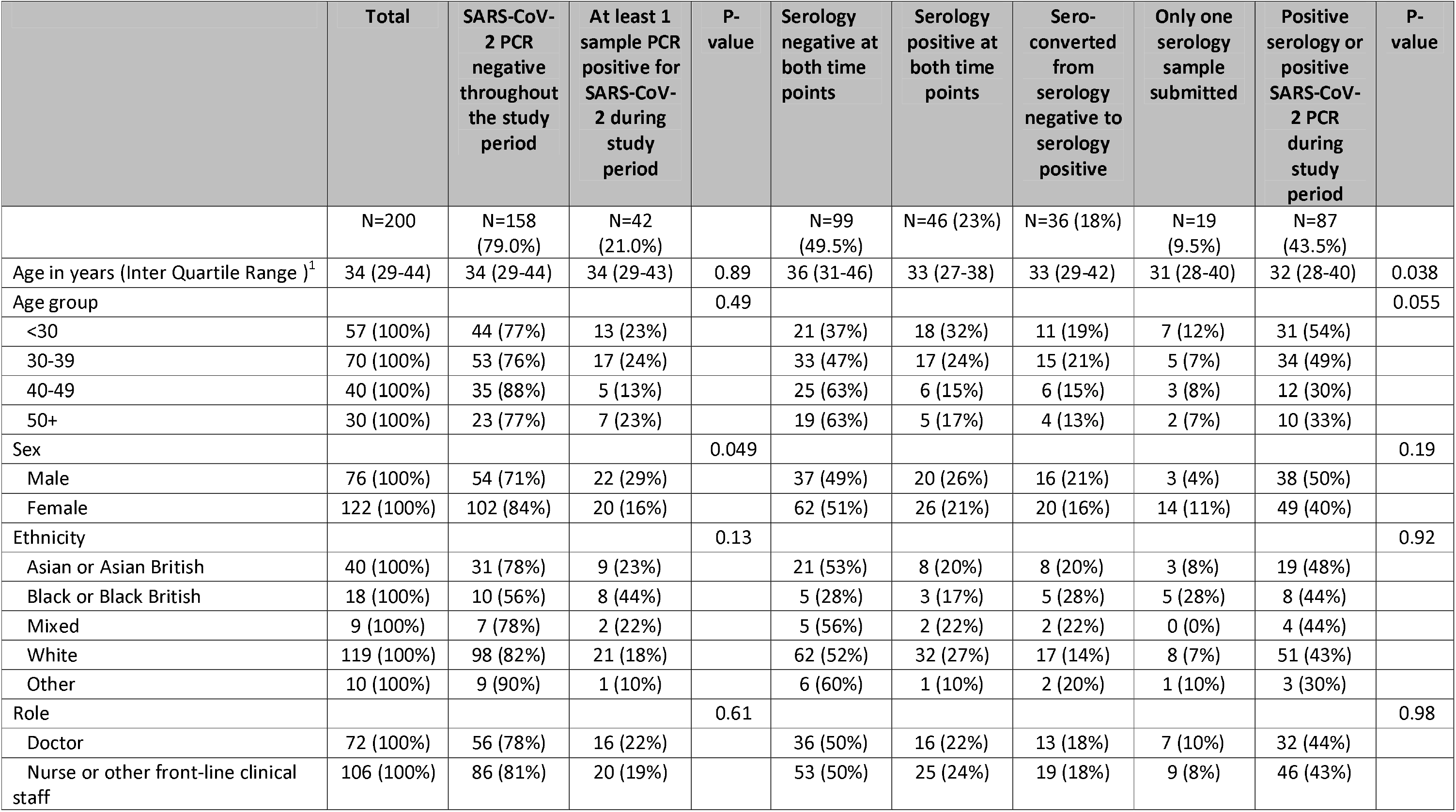
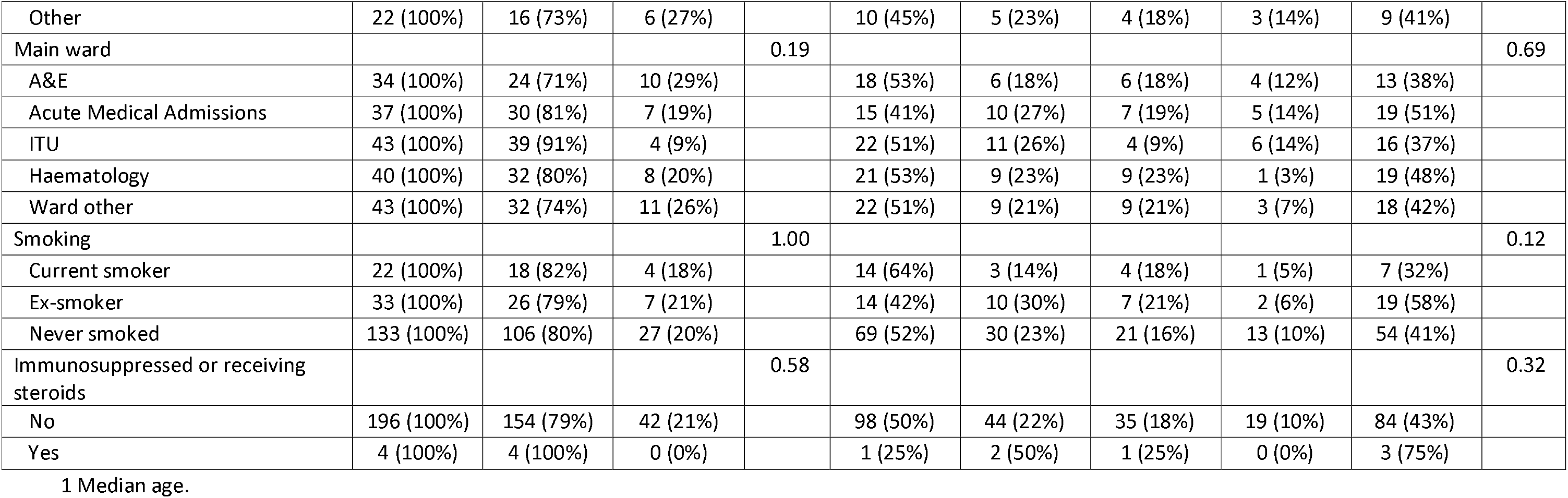
SARS-CoV-2 detection by PCR and by serology in front line health care workers

During the first month of the study, 42 of the 200 HCWs (21.0%, 95% confidence interval (CI) 15.9-27.3) provided at least 1 self-administered swab which was PCR positive for SARS-CoV-2. A greater proportion of men tested positive 22/76 (29.0%, 95% CI 19.7-40.4) than women 20/122 (16%, 95% CI 10.8-24.2) p=0.0498. There was no strong evidence of a difference by area of work in the proportion who returned a positive swab.

Using the composite serological results described, 82/181 (45.3%) HCWs who provided two blood samples were seropositive after one month. This comprised 36 (19.9%) who seroconverted during the study and 46 (25.4%) who were seropositive at both time points.

87 of 200 HCW (43.5%) had evidence of SARS-CoV-2 infection; either positive serology, or RNA detection at any time-point. Evidence of infection was associated with being under 30 years old (31/57 positive, 54.4%) compared to being over 50 years (10/30 positive; 33.3%).

The median duration of detection of SARS-CoV-2 by PCR during follow up was 8 days, IQR 5-12. The longest observed duration of SARS-CoV-2 detection was 22 days. Participants’ time in the study is illustrated in Figure 1.

**Figure 1.**
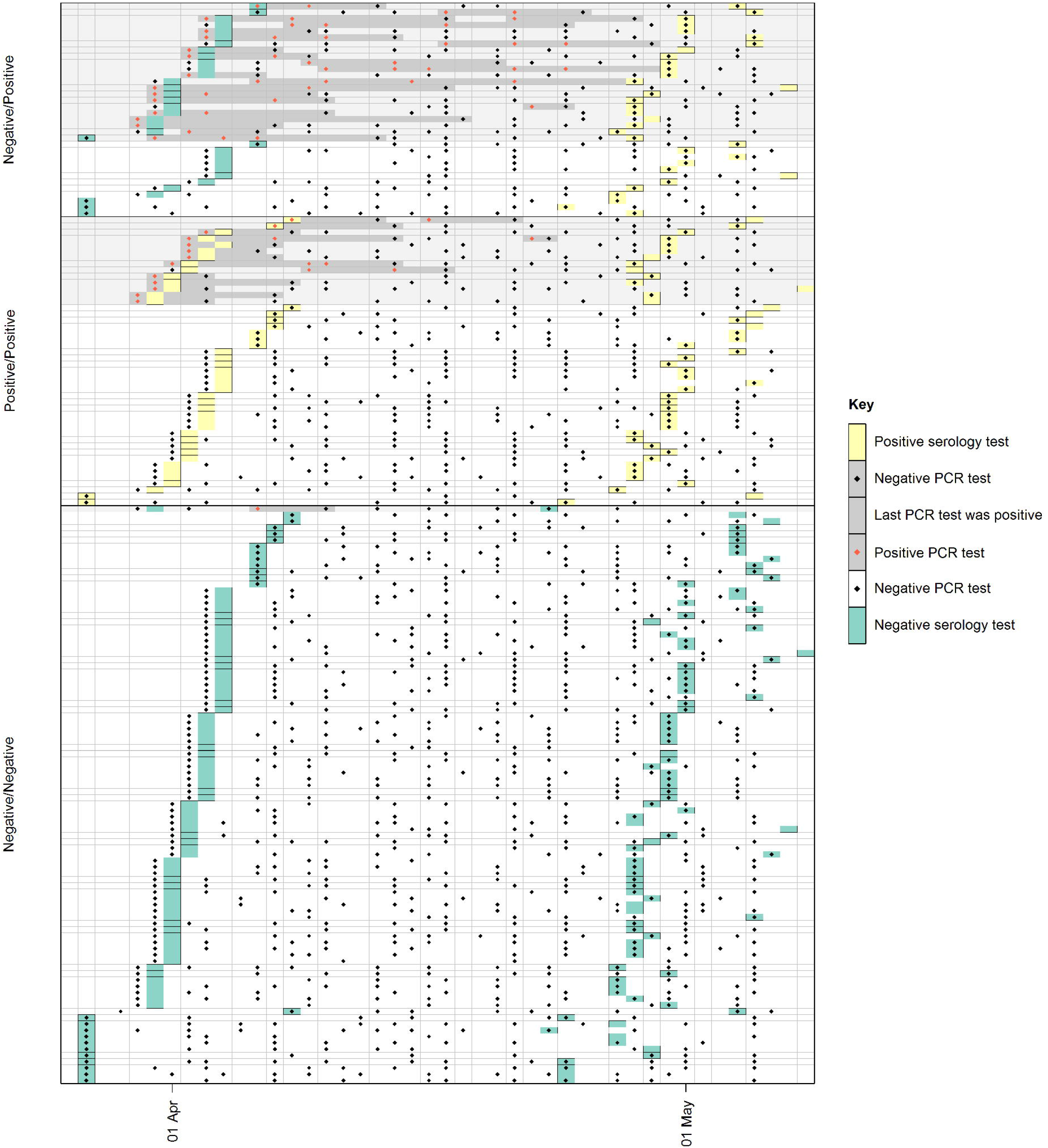
Detection of SARS-CoV-2 by PCR in nose and throat swabs and by serology from 200 front line health care workers

To assess the potential protective effect of antibodies to spike-protein on subsequent development of PCR confirmed infection we compared the risk of PCR positive disease in the one month of follow up in those who were serology negative and PCR negative at baseline, with those who were serology positive and PCR negative at baseline. We excluded 12 of the 120 HCWs who were serology negative and PCR negative at baseline who seroconverted without having had a positive swab during follow (these may represent seroconversions from infections acquired before the baseline sample). Of the remaining 108, 99 remained PCR negative, 9 became PCR positive and seroconverted and 1 became PCR positive but had not seroconverted to date (in a blood sample taken 17 days later). This represents a 9.3% (10/108) infection rate over the one month of follow up in those with no evidence of antibodies or viral shedding at baseline. In 13 of the 46 HCWs with positive serology at enrollment, SARS-CoV-2 RNA was also detected by PCR around the same time. Of 33 staff who had positive serology but were PCR negative at enrolment, 32 remained PCR negative through follow up. One HCW had a PCR positive swab six days later, despite a negative PCR test at enrollment, which likely represented a waning infection.

26/200 HCW were PCR positive at baseline (13%). Of these, 13 (50%) were already antibody positive at baseline and the remaining 13 (50%) had seroconverted by the 1 month follow up sample.

All of the 46 HCW with positive antibodies at baseline had positive antibodies in their follow up samples approximately one month later.

Of the 36 who seroconverted during the study, 19 had SARS-CoV-2 RNA detected either at the time of enrolment (when seronegative) or in the 7 days following enrolment. Of the 36 seroconversions during the study, 12 were in staff in whom no SARS-CoV-2 was detected by PCR during follow-up. Of the 99/200 who were seronegative at both time points, only one tested PCR positive (on a single swab taken 17 days before their second serology test).

Only 8/42 HCW (19%) who were PCR positive during the study period had symptoms that met the current case definition of cough or fever or altered sense of taste or smell (table 2). This represents 8/13 symptomatic cases (61%). Expanding this to those who reported symptoms in the 2 weeks around the time of a PCR positive sample during follow up, only 2/16 (13%) had symptoms compatible with current case definition. No participants had an illness severe enough to require hospital admission. Over time, SARS-CoV-2 PCR Ct values increased, in keeping with reducing RNA detection, although PCR was still positive in 17/42 after day 8 (supplementary figure 1).

**Table 2.** Reported symptoms and SARS-CoV-2 detection by PCR or serology in front-line health care workers

|  | All participants | PCR |  |  | Serology |  |  |  |
| --- | --- | --- | --- | --- | --- | --- | --- | --- |
|  |  | All tests negative<sup>1</sup> | 1+ test positive<sup>2</sup> | New positives, with 2-week window<sup>3</sup> | Negative / Negative<sup>4</sup> | Positive / Positive<sup>5</sup> | Sero-converted<sup>6</sup> | Other |
| All participants | 200 | 158 | 42 | 16 | 99 | 46 | 36 | 19 |
| Asymptomatic | 155 (78%) | 128 (81%) | 27 (64%) | 6 (38%) | 80 (81%) | 37 (80%) | 21 (58%) | 17 (89%) |
| Any symptoms | 45 (23%) | 30 (19%) | 15 (36%) | 10 (63%) | 19 (19%) | 9 (20%) | 15 (42%) | 2 (11%) |
| Cough or fever or anosmia | 21 (11%) | 13 (8%) | 8 (19%) | 2 (13%) | 7 (7%) | 9 (20%) | 4 (11%) | 1 (5%) |
| Runny nose | 22 | 14 | 8 | 4 | 10 | 2 | 9 | 1 |
| Sore throat | 16 | 11 | 5 | 1 | 5 | 5 | 4 | 2 |
| Headache | 14 | 9 | 5 | 2 | 4 | 3 | 6 | 1 |
| Cough | 10 | 6 | 4 | 1 | 3 | 5 | 2 | 0 |
| Fatigue | 9 | 5 | 4 | 2 | 2 | 2 | 4 | 1 |
| Change in sense of taste or smell | 9 | 4 | 5 | 1 | 1 | 4 | 4 | 0 |
| Muscle aches | 8 | 4 | 4 | 2 | 0 | 2 | 5 | 1 |
| Difficulty in breathing | 6 | 5 | 1 | 1 | 3 | 2 | 0 | 1 |
| Other symptoms | 5 | 2 | 3 | 3 | 2 | 0 | 3 | 0 |
| Fever | 4 | 2 | 2 | 1 | 1 | 1 | 2 | 0 |
| Joint pain | 4 | 4 | 0 | 0 | 1 | 1 | 1 | 1 |
| Chest pain | 1 | 1 | 0 | 0 | 0 | 1 | 0 | 0 |

1. HCWs were negative for SARS-CoV-2 by PCR in all nose and throat swab samples submitted. 2. HCWs who had at least 1 samples positive for SARS-CoV-2 by PCR. 3. HCWs who were positive by PCR for SARS-CoV-2 excluding those PCR positive at baseline AND who reported symptoms in the two weeks before their first, after their last or during their positive swabs. 4. HCWs in whom serology tests for SARS-CoV-2 were negative at enrolment and follow up. 5. HCWs who were sero-positive at both enrolment and follow up. 6. HCWs who were serology negative at study enrolment and serology positive at the follow up point. 7. HCWs who did not submit two serology samples

In this cohort of HCWs, the week in which most cases occurred was 30th March-5th April which coincided with the highest number of new cases in London (Figure 2).

**Figure 2.**
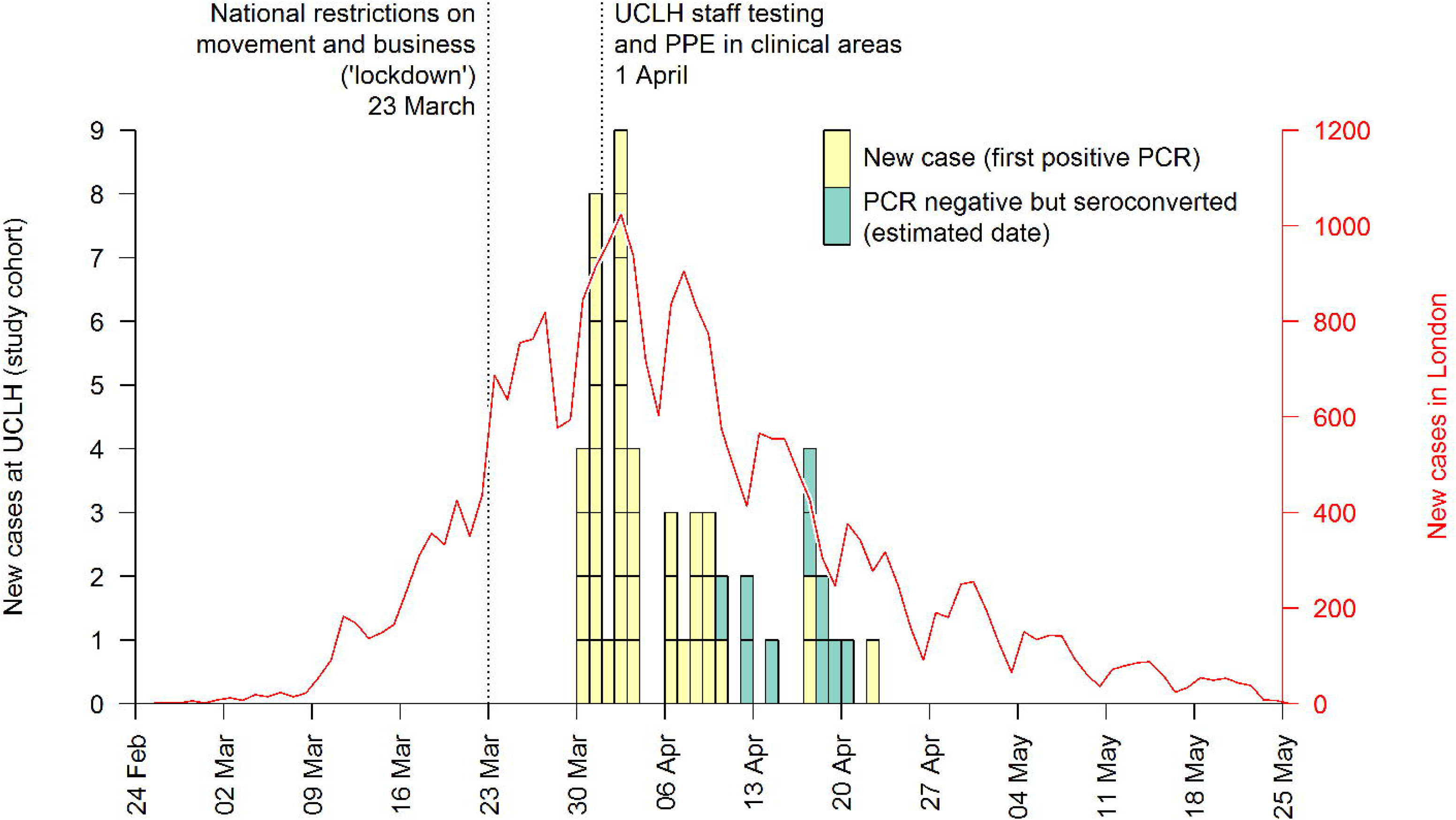
Epidemiological curve of daily confirmed cases of SARS-CoV-2 in front line health care workers at the hospital, and reported cases in London. PCR negative participants who seroconverted are shown at the midpoint between baseline and follow-up serology tests.

## Discussion

We present results for the first month of a cohort study involving 200 front line HCW who were intensively followed with serology and nose and throat swabs during the peak of the COVID-19 pandemic in central London, and report a high proportion with evidence of SARS-CoV-2 infection. Almost half (45%) of front line HCWs who worked in areas with the highest burden of COVID-19 patients (ICU, AMU, A&E, COVID cohort wards) had acquired SARS-CoV-2 by early May 2020. We also show that all but one HCW with PCR evidence of infection developed antibodies; the single HCW who was seronegative had antibodies checked 17 days after their positive swab so may not have had sufficient time to seroconvert. Antibodies to the spike protein of SARS-CoV-2 remained detectable over the one month of follow up. There was no evidence of reinfection in those with antibodies at baseline over one month follow up. However, the number was small and longer follow-up is essential. We emphasised the important message that those with evidence of seropositivity should not alter protective behaviours.

Other studies have also found a high level of infection in HCWs. A cross sectional study combining SARS-CoV-2 seroprevalence with nose and throat swab PCR in 578 randomly selected HCWs in a large teaching hospital in Barcelona found a combined point prevalence of infection of 11% (16). Our study shows a cumulative prevalence of 43.5%. In the UK, a study in Birmingham, conducted at a similar time to ours, found 24% of 516 HCWs had serological evidence of SARS-CoV-2 at the end of April 2020 (4); although the study showed a higher proportion of seropositive HCWs in acute and general medicine (33%), this was still lower than the 45% seen in our cohort. HCWs in both cities were following similar personal protective equipment guidance. At the time of writing, London has had the highest numbers of COVID-19 hospitalisations and deaths in the country, which may account for the higher infection levels seen in London HCWs. Seroprevalence studies conducted by Public Health England show that around 15% of the general public in London were seropositive in week 18, which coincides with the second serology testing in our cohort, by which time 45% of the HCWs were seropositive (17). This suggests that the majority of the infection in HCWs is due to occupational exposure, however it should also be noted that a high proportion of the general London population were locked down during this period so it is possible some of the excess risk in health care workers may be due to non-work exposures, for example during travel to work which often includes use of the underground with crowding and heavy contact with potentially contaminated surfaces. The high levels of infection seen in London HCWs highlights the risk to this group of acquiring COVID-19 in their workplace and supports a recent government move to recommend asymptomatic screening for all HCWs, as we previously advocated (18).

In our cohort, a higher proportion of men were PCR positive than women while serological evidence of infection was similar in both groups. A general population survey of SARS CoV-2 in nose and throat swabs in the general UK population (the ONS study) shows similar prevalence in men and women (19). Men however, are known to have a much higher mortality rate; the mechanisms underlying this require further investigation (17). Although being male is associated with a poor outcome from COVID-19 (18), no health care workers in our cohort were admitted to hospital with severe illness. A higher proportion of those under 30 years old had evidence of SARS-CoV-2 infection than in those over 50. This work is descriptive and we do not include multivariate or adjusted analysis since the numbers were too small to develop robust multivariate models. However, this difference may reflect a difference in behaviour relating to PPE, time-spent in direct patient contact, or performing high risk procedures and further investigation with a behavioural study is being undertaken.

In our study, there was one individual who had not seroconverted by 17 days after a single positive SARS-CoV-2 PCR on a nose and throat swab. This is inconsistent with the documented sensitivity of the serology assays used for this study (flow cytometry detects anti-S IgG IgA and IgM with a sensitivity of 98.8% and ELISA anti-S1 IgG has a sensitivity of 92.1% after day 17) (13). It is possible that the swab was a false positive although negative controls were included in the pipeline and all samples run in duplicate (11). The participant is under follow up. HCWs who were seropositive at enrolment often had detectable SARS-CoV-2 in their swabs indicating possible primary infection around the time of enrolment. There was no evidence that HCWs seropositive at enrolment had a second infection after multiple negative samples. The single seropositive participant with a positive swab 6 days after their enrolment was probably at the end of an infection just prior to enrolment. Our planned longitudinal follow up of seropositive HCWs for at least a year is essential to answer questions about reinfection.

As expected, those with positive tests were more likely to have reported symptoms. Because most of the PCR positive cases in our study were positive at baseline it is possible that participants had symptoms prior to enrolment. Since HCWs were also recruited on the basis of being asymptomatic at baseline this will also bias the estimates of the proportion of cases who are asymptomatic. Our data further support and highlight the known challenge of asymptomatic infection in HCWs(5,7) although the extent of this is difficult to quantify without longer follow up to ensure incident cases are captured. Overall, a small proportion of those who seroconverted or who had a positive PCR reported any symptoms, and in particular symptoms meeting the case definition. Although the numbers are small, of nine participants who reported an alteration to taste and/or smell, eight had a positive serology test. Anosmia and dysgeusia have been associated with SARS-CoV-2 infection in large studies (20) and have been recently added to the case definition in the UK (15). At the time of this study, however, the symptom was not included in the case definition and HCWs were not advised to self-isolate. Such restrictive symptom criteria may have contributed to inadequate isolation and subsequent hospital transmission and highlight the need for strategies employing routine asymptomatic surveillance.

The duration of infection seen in our cohort was a median of 8 days which is shorter than reported(21). Since most HCWs in our cohort were already PCR positive at baseline, viral shedding might be underestimated in our study. As expected, Cycle threshold values in RT-PCR increased over time, in keeping with reducing detection of viral RNA quantity. Although Ct values are not absolutely quantitative and there are no standardised accepted cut-offs in relation to infectivity, strong inverse correlation with the ability to recover infectious virus has been reported(22). In our cohort, the Ct values observed after day 8 were in the range 30-36 with detection up to day 28 with a CT of 35. Further investigation of infectivity of isolated virus is warranted.

HCW infections in our cohort occurred in tandem with the peak seen in London and reduction following the restriction of movement. A further significant change seen in healthcare facilities was the use of enhanced PPE by HCWs in all clinical areas (10). Although our data cannot provide direct evidence of the effectiveness of this intervention, the incidence of new infections following this change in policy was lower than prior to the change. The high levels of infection seen in HCWs is likely to be indicative of transmission from patients to HCWs (which is likely to be minimised by appropriate use of PPE) and transmission from HCW to HCW when PPE is not being worn. The limitations of this study lie in the challenge of following a cohort of individuals who work irregular shifts, in stressful environments, at the peak of the pandemic and often travel to work from long distances. The study aim was to capture infections with twice weekly swabs and avoid missing short lived or transient infections. In order to do this, we sampled HCWs whether they were working a shift or not, providing significant strength to our study. However, gaps in follow-up where participants have not submitted twice-weekly swabs can be seen and there are some who have seroconverted between time points without SARS-CoV-2 RNA being detected. Samples were occasionally held in participants’ home fridges for up to 1 week before they were submitted to the laboratory with potential RNA degradation during these times. This is a preliminary report of the first month of follow up of the London cohort. Future analysis including all study data across study sites will examine the risk by HCW occupation and other demographic variables.

We report early results from a closely observed cohort of front line HCWs who worked throughout the peak of COVID-19 in central London and have the highest seroprevalence in HCWs published to date. This has implications for the expected ‘second wave’ during which hospital admissions are likely to increase again and is applicable to similar large inner-city hospital settings. In light of the rapid launch of vaccine trials, our data can provide useful insights for strategic planning of potential vaccination roll out. Our cohort provides limited early evidence of a lack of re-infection in the first month of follow up in those seropositive, with further data to follow. Finally, the high rate of infection combined with further evidence of asymptomatic carriage of virus highlights the importance of appropriate PPE use and strengthens calls for regular surveillance with swabs in HCWs.

## Data Availability

No external datasets

## Acknowledgements

We thank all of the dedicated health care workers who have worked bravely and tirelessly on the front line and taken the time and effort to contribute their valuable samples and questionnaires to this study

## Author contributions

Designed the study CH EN AH NW

Created the study forms and non-lab SOPs CH NV AMcB NW

Study submission (ethics and funding) CH MS EN AH RG NV NW

Study coordination NV CH AMcB MS EN

Collected the data (laboratory and clinical) JH RM SA LE AS HL AMcB NV, Safer Field Team

Managed the samples JH

Managed the data TBSA

Performed and interpreted serology GK LMcC PC

Performed and interpreted the PCRs DM NF GC

Analysed the data TB CH DL

Generated the figures TB DL

Interpreted the combined data EN AH RG LMcC GK RB CH TB DL MS AMcB DM

Wrote the first draft CH

Commented on the draft and agreed the final publication All

## References

1. WHO. Consensus document on the epidemiology of severe acute respiratory syndrome (SARS). 2003.

2. Chowell G, Abdirizak F, Lee S, et al. Transmission characteristics of MERS and SARS in the healthcare setting: a comparative study. BMC Med. 2015;13:210.

3. Pan A, Liu L, Wang C, Guo H, Hao X, Wang Q, et al. Association of Public Health Interventions With the Epidemiology of the COVID-19 Outbreak in Wuhan, China. JAMA [Internet]. 2020 May 19 [cited 2020 May 25];323(19):1915. Available from: https://jamanetwork.com/journals/jama/fullarticle/2764658

4. Shields AM, Faustini SE, Perez-Toledo M, Jossi S, Aldera E, Allen JD, et al. SARS-CoV-2 seroconversion in health care workers, [cited 2020 May 24]; Available from: https://doi.org/10.1101/2020.05.18.20105197

5. Rivett L, Routledge M, Sparkes D, Warne B, Bartholdson J, Cormie C, et al. Screening of healthcare workers for SARS-CoV-2 highlights the role of asymptomatic carriage in COVID-19 transmission. Elife. 2020;epub ahead of print.

6. Hunter E, Price DA, Murphy E, van der Loeff IS, Baker KF, Lendrem D, et al. First experience of COVID-19 screening of health-care workers in England. Vol. 395, The Lancet. Lancet Publishing Group; 2020. p. e77-8.

7. Treibel TA, Manisty C, Burton M, McKnight Á, Lambourne J, Augusto JB, et al. COVID-19: PCR screening of asymptomatic health-care workers at London hospital. Lancet. 2020 May 23;395(10237): 1608-10.

8. Mizumoto K, Kagaya K, Zarebski A, Chowell G. Estimating the asymptomatic proportion of coronavirus disease 2019 (COVID-19) cases on board the Diamond Princess cruise ship, Yokohama, Japan, 2020. Eurosurveillance [Internet]. 2020 Mar 12 [cited 2020 May 28];25(10):2000180. Available from: https://www.eurosurveillance.org/content/10.2807/1560-7917.ES.2020.25.10.2000180

9. Kimball A, Hatfield KM, Arons M, James A, Taylor J, Spicer K, et al. Asymptomatic and presymptomatic SARS-COV-2 infections in residents of a long-term care skilled nursing facility - King County, Washington, March 2020. Vol. 69, Morbidity and Mortality Weekly Report. Department of Health and Human Services; 2020. p. 377-81.

10. COVID-19: infection prevention and control guidance. 2020.

11. Aitken J, Ambrose K, Barrell S, Beale R, Bineva-Todd G, Biswas D, et al. Scalable and Resilient SARS-CoV2 testing in an Academic Centre, [cited 2020 May 23]; Available from: https://doi.org/10.1101/2020.04.19.20071373

12. Grant PR, Turner MA, Shin GY, Nastouli E, Levett U. Extraction-free COVID-19 (SARS-CoV-2) diagnosis by RT-PCR to increase capacity for national testing programmes during a pandemic. bioRxiv. 2020 Apr 9;19:2020.04.06.028316.

13. Ng K, Faulkner N, Cornish G, Rosa A, Earl C, Wrobel A, et al. Pre-existing and de novo humoral immunity to SARS-CoV-2 in humans. bioRxiv. 2020 May 15;2020.05.14.095414.

14. Harris PA, Taylor R, Thielke R, Payne J, Gonzalez N, Conde JG. Research electronic data capture (REDCap)--a metadata-driven methodology and workflow process for providing translational research informatics support. J Biomed Inform [Internet]. 2009 Apr [cited 2019 Feb 13];42(2):377–81. Available from: http://www.ncbi.nlm.nih.gov/pubmed/18929686

15. COVID-19: investigation and initial clinical management of possible cases - GOV.UK [Internet], [cited 2020 May 29]. Available from: https://www.gov.uk/government/publications/wuhan-novel-coronavirus-initial-investigation-of-possible-cases/investigation-and-initial-clinical-management-of-possible-cases-of-wuhan-novel-coronavirus-wn-cov-infection

16. Garcia-basteiro AL, Moncunill G, Tortajada M, Vidal M, Santano R, Sanz S, et al. Seroprevalence of antibodies against SARS-CoV-2 among health care workers in a large Spanish reference hospital. medRxiv. 2020 May 2;1-36.

17. Public Health England. Weekly Coronavirus Disease 2019 (COVID-19) Surveillance Report. [Internet]. 2020 [cited 2020 Jun 2], Available from: https://assets.publishing.service.gov.uk/government/uploads/system/uploads/attachment_data/file/888254/COVID19_Epidemiological_Summary_w22_Final.pdf

18. Black JRM, Bailey C, Przewrocka J, Dijkstra KK, Swanton C. COVID-19: the case for health-care worker screening to prevent hospital transmission. Vol. 395, The Lancet. Lancet Publishing Group;2020.p. 1418-20.

19. National laboratories [Internet], [cited 2020 Mar 12], Available from: https://www.who.int/emergencies/diseases/novel-coronavirus-2019/technical-guidance/laboratory-guidance

20. Yan CH, Faraji F, Prajapati DP, Boone CE, DeConde AS. Association of chemosensory dysfunction and Covid-19 in patients presenting with influenza-like symptoms. Int Forum Allergy Rhinol [Internet]. 2020 Apr 12 [cited 2020 May 29]; Available from: http://doi.wiley.com/10.1002/alr.22579

21. Sethuraman N, Jeremiah SS, Ryo A. Interpreting Diagnostic Tests for SARS-CoV-2. JAMA - Journal of the American Medical Association. American Medical Association; 2020.

22. Wölfel R, Corman VM, Guggemos W, Seilmaier M, Zange S, Müller MA, et al. Virological assessment of hospitalized patients with COVID-2019. Nature. 2020 Apr 1;581(7809):465–9.

